# Impact of housing conditions on changes in youth’s mental health following the initial national COVID-19 lockdown: A cohort study

**DOI:** 10.1101/2020.12.16.20245191

**Authors:** Jonathan Groot, Amélie Cléo Keller, Andrea Joensen, Tri-Long Nguyen, Anne-Marie Nybo Andersen, Katrine Strandberg-Larsen

**Affiliations:** All authors are affiliated Section of Epidemiology, Faculty of Health and Medical Sciences, University of Copenhagen. Postal address: Øster Farimagsgade 5, bd. 24, PO Box 2099, DK - 1014 Copenhagen K.

**Keywords:** COVID-19, housing, mental health, youth, epidemiology, environmental epidemiology

## Abstract

**Background:** Youth’s mental health has on average declined initially during the pandemic and few studies have investigated whether these declines were dependent on housing conditions.

**Methods:** We used data from 7445 youth from the Danish National Birth Cohort (DNBC), collected at participants’ 18^th^ year of life and subsequently three weeks into the initial national lockdown (April 2020). We examined associations between housing conditions (access to outdoor spaces, urbanicity, household density, and household composition) and changes in mental health parameters (mental well-being, Quality of Life (QoL) and loneliness. We report results from multivariate linear and logistic regression models.

**Findings:** Youth without access to outdoor spaces had a greater decrease in mental well-being compared to those with a garden, mean difference: -0·83 (95 % CI -1·19,-0·48), and correspondingly greater odds of onset of low mental well-being, OR: 1·68 (95 % CI 1·15, 2·47). Youth in higher density households and those living alone also had greater odds of onset of low mental well-being (OR: 1·23 (95 % CI 1·05, 1·43) and OR: 1·47 (95 % CI 1·05, 2·07), respectively). Onset of low QoL was associated with living in denser households, as well as living alone. Living alone more than doubled odds of onset of loneliness, OR: 2·12 (95 % CI 1·59, 2·82).

**Interpretation:** Not all youth were equally affected by the pandemic and our findings inform policy makers that youth living alone, in denser households, and without direct access to outdoor spaces are especially vulnerable to mental health declines.

**Research in context:** *Evidence before this study:* Mental health is associated with certain housing characteristics, such as access to green space and household composition. Additionally, we know that mental health amongst youth has been especially impacted by the COVID-19 pandemic and/or social restrictions, at a time where a majority of youth spend more time at home. Cross-sectional studies have indicated that housing conditions during the initial lockdowns were associated with mental health among youth.

*Added value to this study:* We are able to provide evidence that housing conditions have been important factors in how youth’s mental health has changed, due to data collections in our cohort before and during the pandemic. We demonstrate that living alone without access to outdoor spaces and in denser households during lockdown are all associated with deteriorations in mental health in a longitudinal design. The deteriorations in mental well-being are at a level indicative of anxiety and/or depression, indicating that these mental health changes are meaningful from a public health perspective. To our knowledge, this is the first study to examine associations longitudinally in a youth cohort.

*Implications of all the available evidence:* Not all youth will be equally affected by the pandemic and social restrictions. Public health recommendations could be that youth avoid living alone, in dense households and without access to outdoor spaces during a lockdown, if this is at all possible to choose. Additionally, mental health and public health professionals should be aware of these vulnerabilities as they seek to assist youth at times when social restrictions are in place to control community transmission. Additionally, as we look to the future and work towards equitable and health-promoting housing, we must consider aspects that are important to mental health during pandemics and otherwise.

## Introduction

Differences in housing conditions may become increasingly apparent when a large portion of the population is mandated or recommended to spend the majority of their time at home, resulting from nationwide lockdowns implemented globally to contain the spread of SARS-CoV-2 in the COVID-19 pandemic. The COVID-19 pandemic has brought with it public health and political actions varying across countries in their degrees of restrictions,^1^ but in all scenarios likely resulting in more time spent in individuals’ residency. Secondary lockdowns have already occurred in some nations and will likely occur in hotspots, where restrictive measures may be chosen to curb national and regional spread. In Denmark, the national lockdown in spring 2020 was announced March 11^th^ and effective from March 13^th^, 2020.^1^ It involved closing of the national boarders, restaurants/bars, sports facilities, schools and public workplaces, and recommendations to social distance and engage in only essential activities.^2^

Previous studies have documented changes in mental health from before to during initial national lockdowns,^3–6^ highlighting how youth have been disproportionately affected.^3,7,8^ A few studies have described cross-sectional associations between housing conditions and mental health during initial lockdowns,^9–11^ as well as before and after measures of loneliness in separate samples.^12^ To our knowledge, no previous studies have documented changes in young people’s mental health from before to during a lockdown in relation to the housing conditions one must ‘stay home’ in. Using prospectively-collected mental health data, we examined whether mental health declines among Danish youth were dependent on their housing conditions during the initial national COVID-19 lockdown.

## Materials and methods

### Population

The Danish National Birth Cohort (DNBC) consists of mothers and offspring from approximately 100 000 pregnancies enrolled in the cohort during the years 1996 to 2002.^13^ The pregnant women responded to computer assisted telephone interviews twice during pregnancy and twice in the child’s two first years of life. Additional follow-ups were conducted at offspring ages 7, 11, and more recently, 18 years. In the 18-year follow up, invitations were sent three months after participants’ 18th birthday. This data collection was initiated in March 2016 and will be completed ultimo 2021; detailed information on DNBC is available at: www.dnbc.dk. Most recently, in the 3^rd^ week of the national Danish COVID-19-related lockdown, mothers and their offspring for whom we had either a private e-mail address or phone number were invited to participate in a COVID-19 online questionnaire, and all participants who responded within a week were re-invited to up to six subsequent consecutive online questionnaires.^2^ In this study, we used data on offspring’s mental health from the 18-year follow-up (*baseline*) as a before lockdown measure and data on housing conditions and mental health from the first of the COVID-19 online questionnaires (*follow-up*) as a during lockdown measure, eFigure 1.

**Figure 1.**
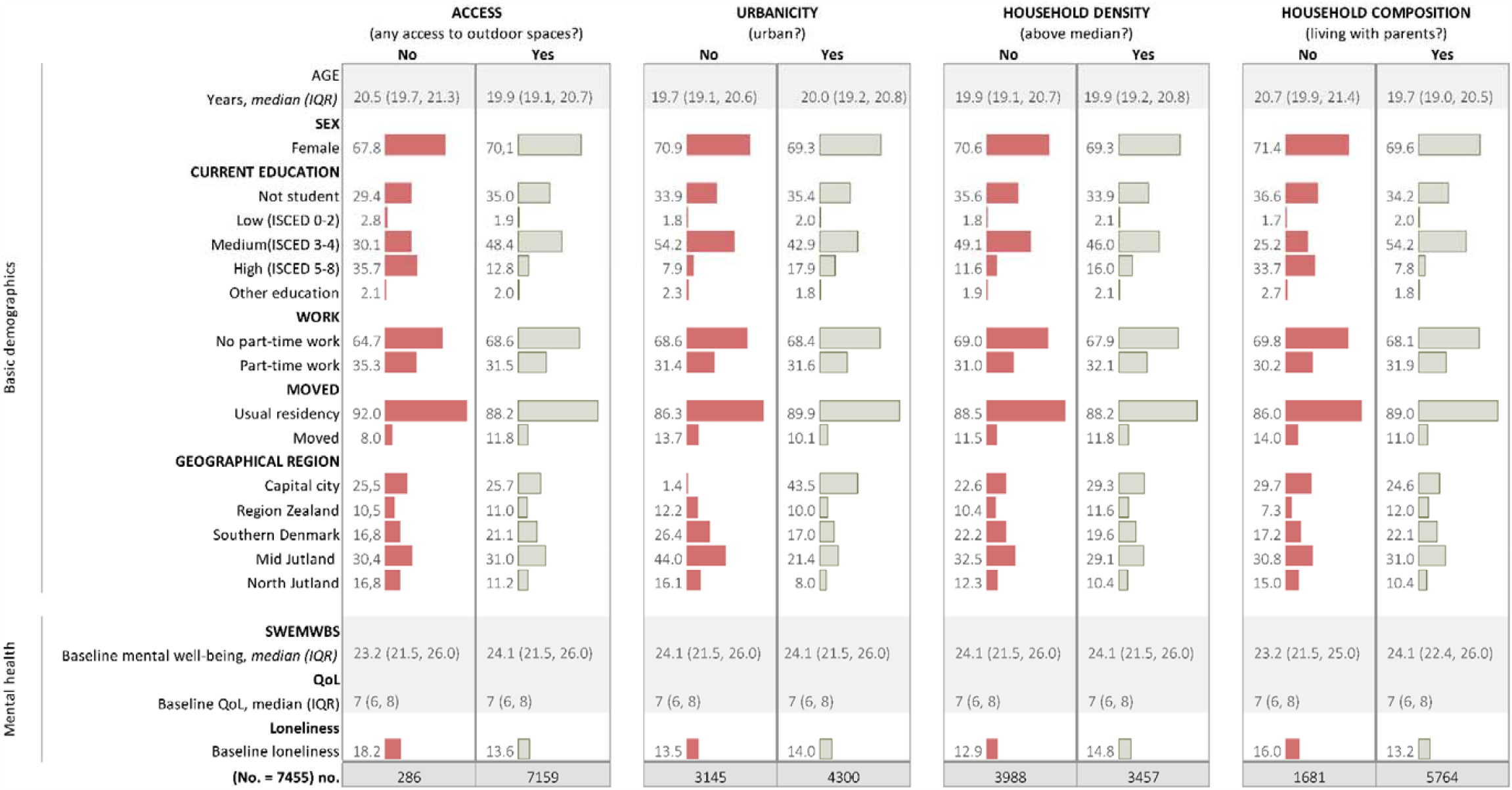
Demographic characteristics by housing conditions (No. = 7445) IQR: Interquartile range, ISCED: International Standard Classification of Education, SWEMWBS: Short Warwick-Edinburgh Mental Well-Being Scale, QoL: Quality of Life. Relative frequencies presented, unless otherwise stated.

### Mental Health Outcomes

We examined changes in mental health with the following three parameters:

#### 1) Mental well-being

Changes in mental well-being were measured as the difference in metric scores on the 7-item Short Warwick-Edinburgh Mental Well-Being Scale (SWEMWBS)^14–16^. This instrument has been validated in a Danish sample, including participants in the age range we studied.^15^ The scale ranges from 7 to 35, with higher values indicating better well-being. Because a score of ≤20 corresponds to possible or probable anxiety or depression,^16^ we used this cut-off to define *low* vs. *normal* mental wellbeing.

#### 2) Quality of life (QoL)

Changes in quality of life (QoL) were reported on an adaptation of the Cantril Ladder scale from 0 to 10, where 0 represented the ‘worst possible life’ and 10 ‘the best possible life’.^17^ In addition to treating the outcome as a continuous variable, we also categorized participants with a score of 5 or lower as having a *low* QoL and above 5 as *normal* QoL, in line with previous research.^17,18^

#### 3) Loneliness

Changes in loneliness were measured only as a binary outcome. Loneliness was measured with similar Likert scales at baseline and follow-up. At baseline, participants were asked ‘How often do you feel lonely?’ with response options: ‘Never’, ‘Occasionally’, ‘Often’, ‘Very often’ or ‘Unsure’ (excluded). ‘Often’ and ‘Very often’ responses were categorized as *lonely* vs. *not lonely*. At follow-up, participants were asked ‘In the last week, how often have you felt lonely?’ with the following response options: ‘Seldom or not at all (less than 1 day)’, ‘Some or a little (1-2 days)’, ‘Occasionally or often (3-4 days)’ or ‘Most of the time (5-7 days)’. Responses with a minimum of three days were categorized as *lonely* vs. *not lonely*.

### Housing Conditions during lockdown

We examined the following four housing conditions:

#### 1) Direct access to outdoor spaces

Direct access to outdoor spaces was defined as not having direct access to outdoor spaces or having direct access to a common yard only, a balcony only, a garden only, or multiple outdoor spaces/other (e.g. a garden and a balcony). For descriptive analyses only, the variable was dichotomized into no or direct access to any of the aforementioned outdoor spaces.

#### 2) Urbanicity

Urbanicity was defined as residential degree of urbanicity, defined by the European Union, using the following municipality-level population density categorizations: rural (thinly populated), semi-urban (intermediate density) and urban (densely populated).^19^ Postal codes for residency at follow-up were self-reported.

#### 3) Household density

Household density was defined as the reciprocal of average square meters per person in the household, i.e. 1/(square meters/number of person in household). The housing area was reported in 10 square meters intervals, e.g. 50 to 59, and we therefore used the middle value. Children and adults were counted similarly, rather than assigning lower value for children. Exposure was split at the median value to create a binary variable for both the descriptive and regression analyses. The continuous variable was used in models where household density was a covariate.

#### 4) Household composition

Household composition was categorized as living alone, with friends or roommates (including in a dormitory), with a partner, with parents only, or with parents and children or siblings. For descriptive analyses, the variable was dichotomized into living without parents (i.e. alone, with a friend/roommate, and partner) or living with parents (i.e. living with parents only and living with parents and siblings/children).

### Covariates

Data on sex (male, female), current educational enrolment (not a student, ISCED 0-2, ISCED 3-4, ISCED 5-8, and ‘other education’), part-time work (yes, no), moving during lockdown (moved from residency/other postal code: yes, no), geographical regions (Capital City Region, Region Zealand, Region of Southern Denmark, Mid Jutland Region, North Jutland Region), and baseline mental well-being/QoL/loneliness were considered potential covariates.

Three variables were considered *a priori* for interaction analyses: sex, quarantine and self-reported psychiatric illnesses. Quarantine was defined as those who at follow-up reported having been in quarantine or isolation during the last 14 days. Psychiatric illnesses were self-reported and respondents could choose between the following answers: ‘no history of psychiatric illness’, ‘current psychiatric illnesses’, ‘previous psychiatric illnesses’, or ‘unsure’. Youth reporting current psychiatric illness and those who were ‘unsure’ were grouped together (since respondents who were ‘unsure’ resembled those with a current psychiatric illness most), and those with previous illnesses or none were grouped together.

### Statistical analyses

Statistical analyses were performed in Stata 15.0 (StataCorp, Tx, USA).

Absolute and relative frequencies are presented for the dichotomized exposure variables and median and interquartile ranges are presented for continuous covariates. Unadjusted mean changes in mental well-being (SWEMWBS) scores were computed for each category in each exposure. Kernel density plots and bar plots were constructed to visualize the distribution of the mental health parameters at baseline and follow-up.

Crude and adjusted regression models were fitted for linear and logistic regressions. Adjusted Model 1 included potential covariates and Adjusted Model 2 included additional mutual adjustments for housing conditions. Results are presented for Adjusted Model 2 and results of crude and Adjusted Model 1 are presented in the eSupplement. Linear regression models were fitted for the two outcomes on a continuous scale (changes in mental well-being and QoL scores). To also examine the odds of onset of *low* mental well-being, *low* QOL and loneliness, models were fitted for participants who were at risk (only individuals who had *normal* levels at baseline, eFigure 1). In *post hoc* analyses, instead of excluding participants with *low* mental wellbeing, *low* QoL, or *loneliness* at baseline, we estimated relative risk ratios (RRRs) for each of the potential combinations of the binary baseline and follow-up mental health parameter in multinomial logistic regression models with the normal-normal (not lonely-not lonely) as reference outcome-(e.g. for mental wellbeing: *low* at baseline followed by *low* at follow-up vs. *normal* at baseline followed by *normal* at follow-up.

For interaction analyses, linear (mental well-being and QoL) and logistic (loneliness) regression models with and without interaction terms were compared, and differences in fit tested with the likelihood ratio test. For tests indicating potential interaction (p <·2), we fitted stratified or joint effect models, as appropriate.

We performed complete case analyses.

## Results

### Participants

We included a total of 7445 of the 9211 young participants who had responded to the first COVID-19 online questionnaire and the 18-year DNBC follow-up, eFigure 1. The age range at follow up was 18 to 23 with a median age of 20 years. Approximately two thirds of the study population were female. Youth with no access to outdoor spaces were older, more educated, and more often lonely at baseline, but had less often moved compared to youth with access to outdoor space. Additionally, in the capital region more lived in households with density above median, and the other housing conditions likewise differed according to demographic characteristics, Figure 1.

### Declines in mental health during lockdown

Lower mental well-being and QoL and higher levels of loneliness were observed in the third week of the lockdown compared to before the lockdown, with higher proportions of individuals with scores indicative of possible (19·1 % compared to 12·2 %) or probable (2·6 % compared to 1·3 %) depression/anxiety, low QoL (36·6 % compared to 15.6 %) or being lonely (23·1 % compared to 13·8 %), Figure 2.

**Figure 2.**
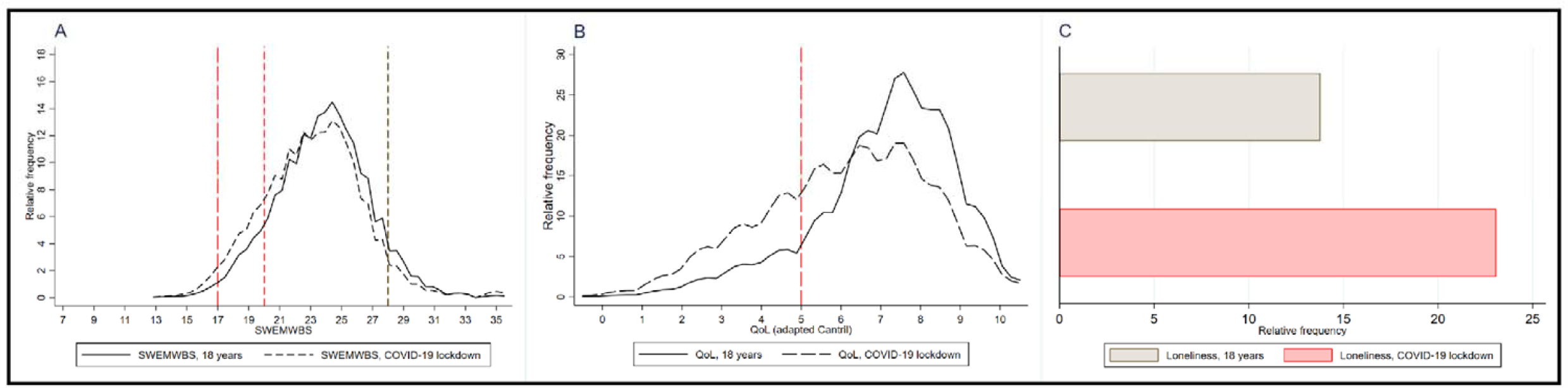
Distribution of baseline and follow-up mental well-being (SWEMWBS) scores, QoL scores, and loneliness (No. = 7445) SWEMWBS: Short Warwick-Edinburgh Mental Well-Being Scale, QoL: Quality of Life. **A)** Kernel density plot of the distribution of mental well-being (SWEMWBS) scores: Long dash red reference for probable anxiety/depression,^16^ short dash red reference line for indication of possible anxiety/depression, and black short reference line for indication of high mental well-being, **B)** Kernel density plot of distribution QoLscores: Long dash red reference for cut-off of low QoL^18^, **C)** Barplot of loneliness. Participants in the tails of SWEMWBS were aggregated in kernel density plots to secure non-identifiability of participants

### Housing conditions impact on changes in mental health

Unadjusted mean changes in mental well-being scores were highest for those with no access to outdoor spaces, Figure 3. For this group the score was >1 point lower, and a 1-point change on the SWEMWBS is considered to represent a clinically meaningful change.^16^ Similarly, in the adjusted model, a lack of direct access to outdoor spaces was associated with the greatest decreases in mental well-being scores (no access vs. garden: mean difference: −0.83 [95 % CI -1.19, −0.48], Figure 4 and eTable 1). A stepwise decrease in odds of onset of low mental well-being was also observed going from more limited to less limited outdoor spaces (Figure 5 and eTable 2). Youth living in denser households had greater decreases in mental well-being (Figure 4 and eTable 1) and likewise greater odds of onset of low mental well-being (Figure 5 and eTable 2) than youth in non-dense households. Household composition was also associated with changes in mental well-being. Youth living alone had greater decreases than those living with parents (Figure 4 and eTable 1). A stepwise greater odds of onset of low mental well-being was observed for youth living with parents and children/siblings, roomies/friends, and alone, compared to living with parents (Figure 5 and eTable 2).

**Figure 3.**
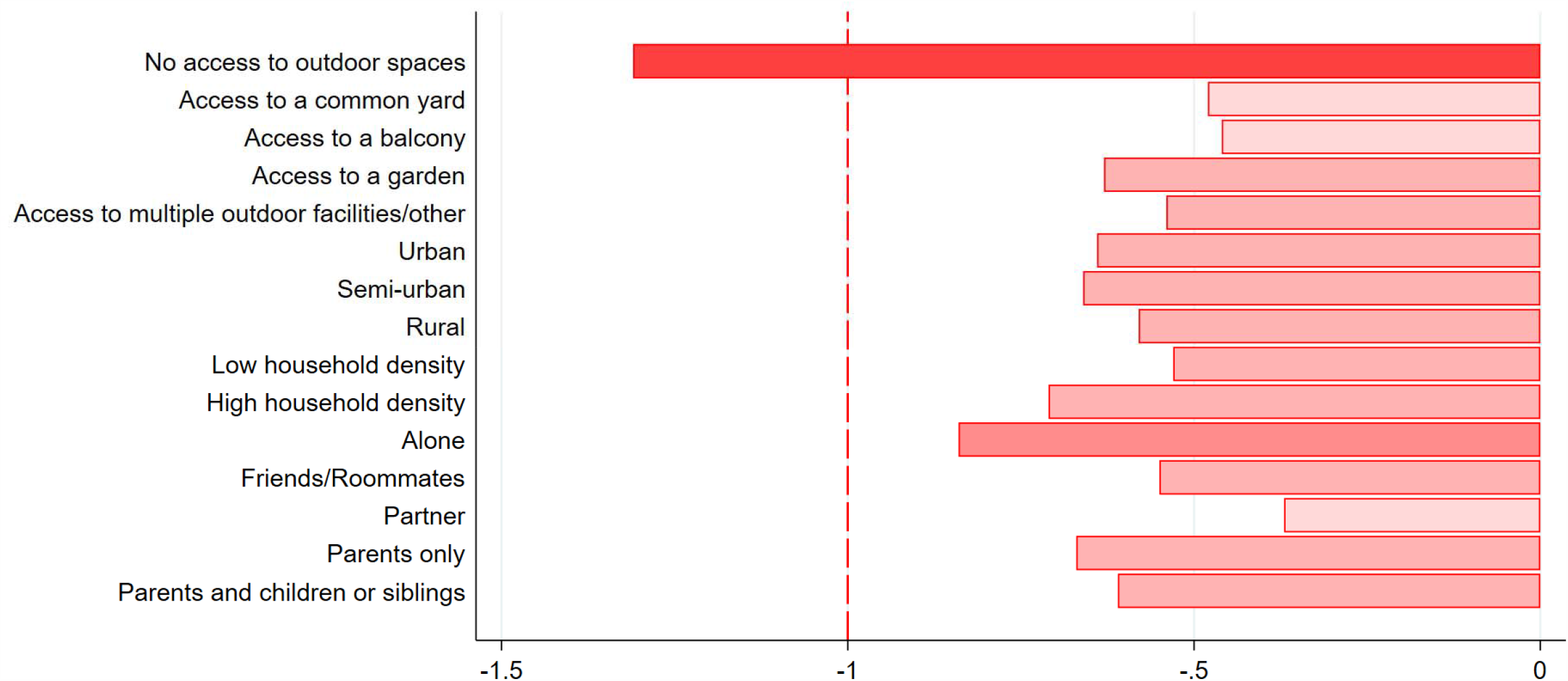
Changes in mental well-being (SWEMWBS) scores indicative of meaningful changes, according to housing conditions (No. =7445) SWEMWBS: Short Warwick-Edinburgh Mental Well-Being Scale Unadjusted means for changes in mental well-being scores on the SWEMWBS, by housing conditions. Red dashed line indicates minimum change considered meaningful at the individual level on the SWEMWBS^16^ and incrementally darker shades of red indicate incrementally larger decreases in SWEMWBS scores.

**Figure 4.**
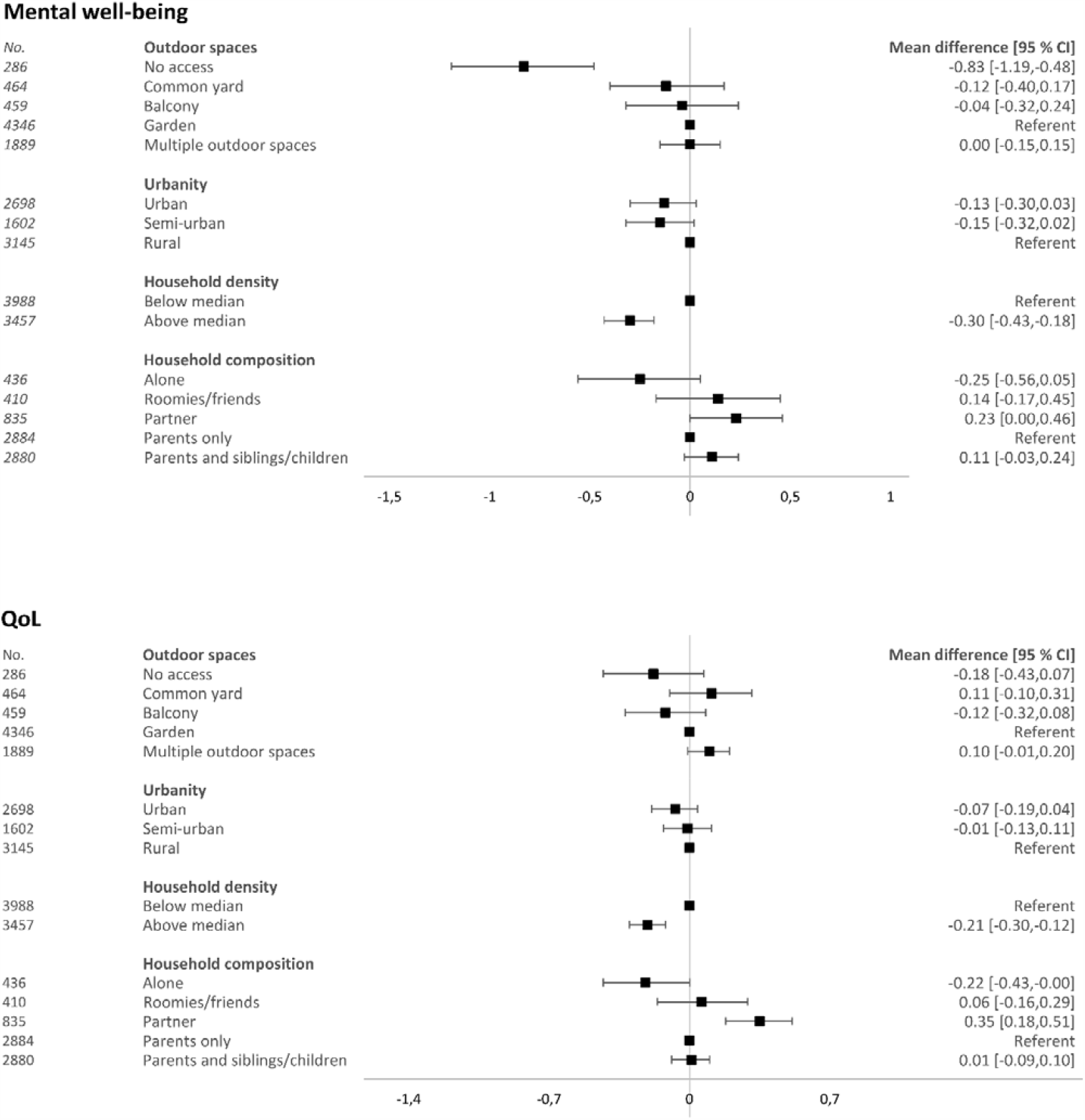
Changes in mental well-being and QoL, according to housing conditions (No. = 7445) QoL: Quality of Life, CI: Confidence Intervals. Mean difference and 95 % CI are presented. Adjusted model 2: adjusted for age, sex, current education, part-time work, moving, geographical region, baseline SWEMWBS/QoL scores, and mutually adjusted for housing conditions

**Figure 5.**
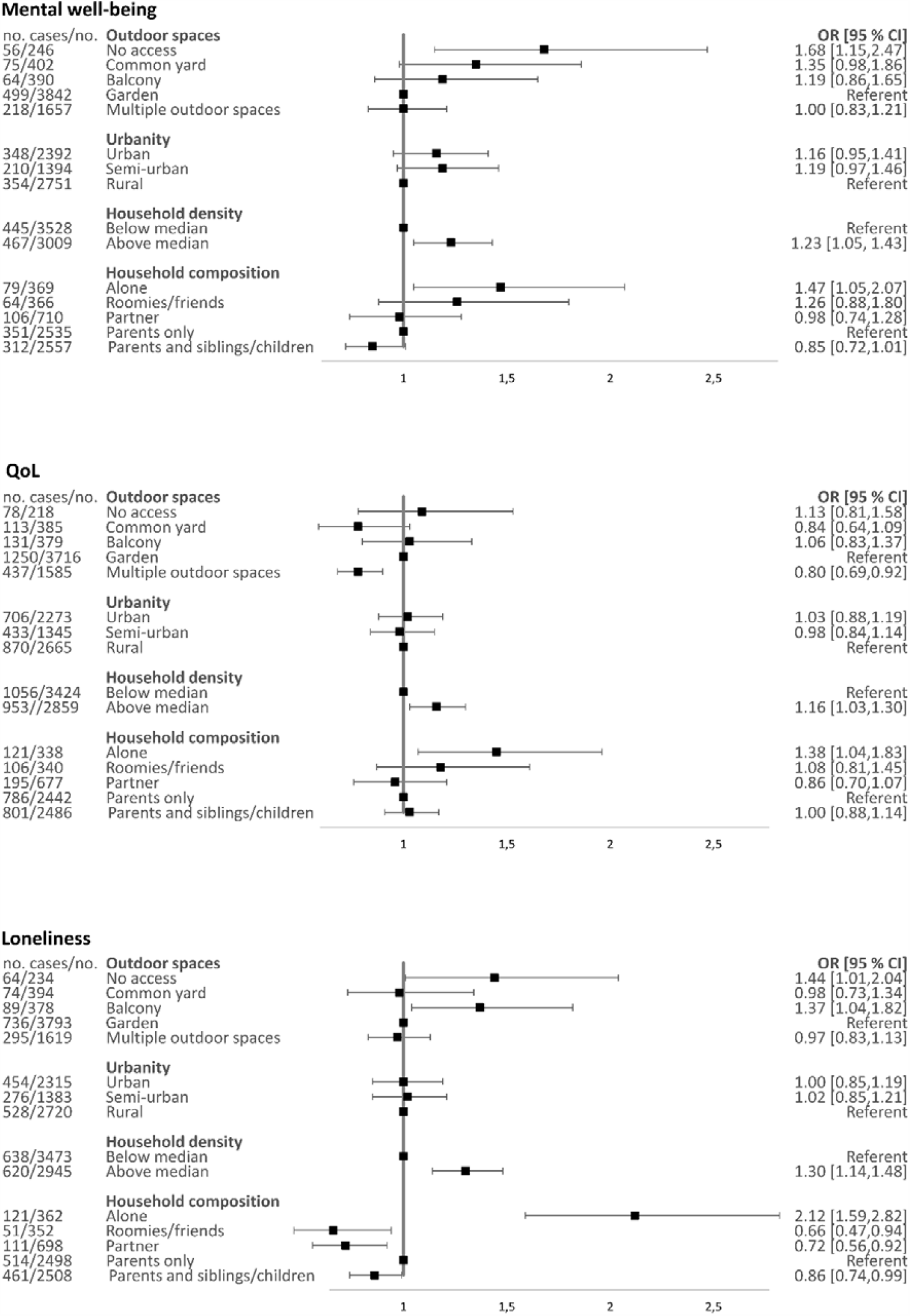
Onsets of low mental well-being, low QoL, and loneliness, according to housing conditions. QoL:Quality of Life, OR: Odds Ratios, CI: Confidence Intervals. OR and 95 % CI are presented. Population at-risk of developing low mental well-being/low QoL/loneliness: Mental well-being (No. = 6537), QoL (No. =6283), Loneliness (No. =6418) Adjusted model 2: adjusted for age, sex, current education, part-time work, moving, geographical region, baseline SWEMWBS/QoL scores/loneliness, and mutually adjusted for housing conditions

Decreases in QoL and onset of low QoL were associated with living in a denser household and living alone (Figure 4 and eTables 3 & 4). Youth living with a partner reported increased QoL (Figure 4 and eTables 3). Incident loneliness was associated with living alone and living in a denser household (OR 2.12 [95 % CI: 1.59, 2.82] and OR 1.30 [95 % CI 1.14, 1.48], respectively: Figure 5 and eTable 5).

Analyses stratified by sex indicated minor sex differences that were not consistent across mental health parameters for household composition (eTable 6). Males experienced greater decreases in mental well-being when living alone or with roomies/friends and in denser households, and greater increases in loneliness when living alone. Females experienced greater decreases in QoL when living alone, but greater increases in QoL when living with their partner. In the joint effects model examining interactions with quarantine status, youth not quarantined and with no direct access to outdoor spaces experienced the greatest decreases in mental well-being (mean difference : −0.98 [95 % CI: -1.38, −0.58], eTable 7). Psychiatric illness did not interact with housing conditions, except for QoL and household composition where youth with no psychiatric illness and living with a partner had increased QoL (0.38 [0.20, 0.55]).

eTable 8).

For the majority of youth for whom the mental health changed from before lockdown it decreased, but for 5.3 % the mental well-being, for 6.1 % QoL and for 7.6 % loneliness improved during the lockdown. Factors associated with onset of low mental well-being, low QoL, or loneliness (no access to outdoor spaces, above median household density, and living alone) were also associated with continued poorer mental health, while these patterns were less clear for improvements in mental health (eTables 9,10,11).

## Discussion

The present study shows that housing conditions during the initial national Covid-19 related lockdown influenced changes in mental health among Danish youth. A lack of access to outdoor spaces was associated with greater decreases in mental well-being. Living in a denser household and living alone were also both associated with decreases in mental well-being and QoL, and with an increase in loneliness. Therefore, characteristics of the built environment and the household matters for the mental health of our youth during this pandemic.

The concepts of ‘a sense of over-crowding in the home’ and ‘escape facilities’ have previously been highlighted as important aspects of the built environment that may influence mental health.^20^ Results from the first studies on housing and mental health in the COVID-19 pandemic, suggest that these factors may be associated with mental health during a pandemic as well.^9–11^ Three cross-sectional studies conducted in Southern European countries which were severely impacted by the pandemic have assessed associations between some housing conditions on mental health during the initial Covid-19 related lockdowns. In Italy, students with moderate–severe and severe depressive symptoms lived in significantly smaller apartments without access to outdoor space, such as a balcony or a garden, and had a poor quality view.^10^ Another study from France found that living in an urban area, having access to an outdoor space and a bigger dwelling size was positively associated with well-being (measured using the WEMWBS scores).^11^ A third study conducted in Portugal reported that access to a garden was related with lower depression and stress, whereas the number of people in the household was not.^9^ In addition, a prospective study in the UK reported that being a young adult, urbanicity, and living alone were associated with greater loneliness – these results were based on a sample during the pandemic and compared levels to another sample with measures prior to the pandemic.^12^ These studies were not, as we did, able to assess changes in mental health parameters from before to during lockdown in the same individuals, and three of them were not specific to youth,^9,11,12^ but their findings are consistent with ours, and highlight the importance of aspects of the built environment previously known to impact mental health.

Although changes in mental health from before to during lockdown differed for females and males, only minor and inconsistent differences in the impact of housing were observed. For example, living with a partner was only associated with higher QoL among females and was only associated with higher mental well-being among males.

Associations between access to outdoor spaces and declines in mental well-being were stronger for youth in quarantine. Given previous findings on the disproportionately negative effects of pandemics on mental health among youth in quarantine,^21^ this may seem counter-intuitive. Strict adherence to quarantine might have an effect that cannot be improved simply by access to outdoor spaces. Alternatively, youth may utilize some of the outdoor spaces investigated, only when they are not in quarantine (for example, by visiting with friends in a common yard).

We suspected that youth with psychiatric illnesses would be more susceptible to the negative effects of the lockdown and that housing conditions could potentially be more strongly associated with mental health outcomes.^6^ We only observed differences for QoL and household composition. Although youth with psychiatric illnesses in our sample have experienced greater declines in mental health on average (for further information visit coronaminds.ku.dk), our results do not suggest that these interact substantially with housing conditions. Nevertheless, it is difficult to draw definitive conclusions, due to the low numbers.

### Strengths and limitations

One of the main strengths of our study design is the longitudinal design with individual-level data on a large sample of youth with measures from before and during the most restrictive phases of the Danish lockdown in spring 2020. The early lockdown of Denmark effectively curbed the spread of COVID-19 and the number of deaths due to COVID-19 were low compared to other European countries at the time of our follow-up.^2^ Therefore, it is less likely that changes in mental health outcomes are due to the pandemic, as opposed to the lockdown. To our knowledge, no other studies have considered longitudinally the effect of housing conditions on youth’s mental health development before and during the Covid-19 pandemic.

Interpretation of our results deserves consideration of a few limitations. The baseline data were collected at age 18 years and three months for all participants, whereas the participants’ ages varied during the lockdown. Thus, the time elapsed between baseline and follow-up was greater for older participants. All analyses were adjusted for age, and we thereby indirectly accounted for different time between baseline and follow up. For older participants, changes in mental health parameters may be underestimated, since their baseline measures represent a younger age than the follow-up measures, and on average reporting on mental health instruments improves by age. Therefore, our estimates could be underestimated, and thereby conservative, in particular for the housing conditions where age is unevenly distributed as for access to outdoor spaces and household composition. Additionally, although we adjusted for what we consider likely to be the most important confounding variables, we cannot exclude the possibility of unmeasured and residual confounding. Access to outdoor spaces could be a proxy for other beneficial aspects of the built environment. Nonetheless, we suspect that such factors would be captured in the model (geographical region, urbanicity, household density) and would likely not have independent effects on changes in mental health.

Misclassification of housing conditions may have influenced our results. Postal codes were used to identify municipality-level degrees of urbanization. However, in a few instances postal codes may be assigned to multiple municipalities and even regions. Nevertheless, the numbers of potential misclassification due to this are minimal.

A final important limitation to consider is the selection into the study. Less than 10% of the young people still enrolled in the DNBC contributed to this study, as a prerequisite was that they were old enough to have 18-year data and participated in the corona survey. Maternally reported household socio-occupational status collected when pregnant, prenatal smoking, maternal age and parity are all predictors for participation. It is likely that these and other factors influenced both participation and mental health, and we presume this would, if anything, have biased our results towards no association. The external generalizability of these findings must be considered in light of the predominantly female participation and the Danish context. In our study population, approximately 60 % of the DNBC offspring 18 and 19 years old were living with parents at the time of responding to the online questionnaire. The vital statistics for the Danish population in 2016 indicate that this is a lower percentage than in the general population, although this may be due to a greater proportion of females, who tend to move away from home earlier.^22^ In other countries, ages for moving away from the parental home is later,^23^ and the relationship between housing characteristics and youth’s mental health under a pandemic might be different. In our study, 12 % had moved during lockdown, and those who moved more often stayed in non-urban municipalities, had access to outdoor spaces and lived with parents. Additionally, we suspect that the effect of housing conditions on changes in mental health are likely to be more pronounced in countries with more severe and lengthy restrictions.^1^ Future studies may elucidate this.

## Conclusion

Youth’s mental health has declined during the initial stage of the COVID-19 pandemic-with some youth especially vulnerable to mental health declines. Living without access to outdoor spaces, alone, or in denser households may increase onset of depression, anxiety, and loneliness. Housing conditions should be emphasized in efforts to keep youth’s mental health intact and public health professionals and policy-makers should consider the increased vulnerability of youth living alone, in denser households, and without access to outdoors spaces.

## Supporting information

eSupplement

## Data Availability

N/A

## Acknowledgment

The Danish National Birth Cohort (DNBC) was established with a significant grant from the Danish National Research Foundation. Additional support was obtained from the Danish Regional Committees, the Pharmacy Foundation, the Egmont Foundation, the March of Dimes Birth Defects Foundation, the Health Foundation and other minor grants. The DNBC Biobank has been supported by the Novo Nordisk Foundation and the Lundbeck Foundation. Follow-up of mothers and children has been supported by the Danish Medical Research Council (SSVF 0646, 271-08-0839/06-066023, O602-01042B, 0602-02738B), the Lundbeck Foundation (195/04, R100-A9193), The Innovation Fund Denmark 0603-00294B (09-067124), the Nordea Foundation (02-2013-2014), Aarhus Ideas (AU R9-A959-13-S804), a University of Copenhagen Strategic Grant (IFSV 2012) and the Danish Council for Independent Research (DFF – 4183-00594 and DFF – 4183-00152). Follow-up of mother and children in the COVID-19 data collection was supported by a grant from the Velux Foundation (grant number 36336).

