## Supplementary material for "Impact of housing conditions on changes in youth’s mental health following the initial national COVID-19 lockdown: A cohort study": eSupplement

**Table of Contents**, page 1

### eTables

**eTable 1.** Changes in mental well-being (No. = 7445), page 2

**eTable 2.** Onset of low mental well-being (No. = 6537), page 3

**eTable 3.** Changes in quality of life (No. = 7445), page 4

**eTable 4.** Onset of low quality of life (No. = 6283), page 5

**eTable 5.** Onset of loneliness (No. = 6418), page 6

**eTable 6.** Changes in mental well-being, quality of life and loneliness, stratified by sex (No. = 7445), page 7

**eTable 7.** Joint effects of housing conditions and quarantine on changes in mental well-being, quality of life and loneliness (No. = 7418), page 8

**eTable 8.** Changes in quality of life, stratified by psychiatric illnesses (No. = 7418), page 9

**eTable 9.** Changes in category of mental well-being (No. = 7445), page 10

**eTable 10.** Changes in category of quality of life (No. = 7445), page 11

**eTable 11.** Changes in category of loneliness (No. = 7445), page 12

### eFigures

**eFigure 1.** Flow of participation, page 13

**eTable 1.** Changes in mental well-being (No. = 7445)

| | | Mean difference (95 % CI) | | |
|---|---|---|---|---|
| | *no.* | **Crude Model** | **Adjusted model 1** | **Adjusted model 2** |
| **Direct access to outdoor spaces** | | | | |
| None | *286* | -0.68 [-1.03,-0.33] | -0.91 [-1.24,-0.59] | -0.83 [-1.19,-0.48] |
| Common yard | *464* | 0.15 [-0.13,0.44] | -0.14 [-0.41,0.12] | -0.12 [-0.40,0.17] |
| Balcony | *459* | 0.17 [-0.11,0.45] | -0.07 [-0.33,0.20] | -0.04 [-0.32,0.24] |
| Garden | *4346* | 0.00 [0.00,0.00] | 0.00 [0.00,0.00] | 0.00 [0.00,0.00] |
| Multiple outdoor spaces | *1889* | 0.09 [-0.07,0.25] | -0.00 [-0.15,0.14] | 0.00 [-0.15,0.15] |
| **Urban** | | | | |
| Urban | *2698* | -0.05 [-0.21,0.10] | -0.16 [-0.32,0.00] | -0.13 [-0.30,0.03] |
| Semi-urban | *1602* | -0.08 [-0.25,0.10] | -0.16 [-0.33,0.02] | -0.15 [-0.32,0.02] |
| Rural | *3145* | 0.00 [0.00,0.00] | 0.00 [0.00,0.00] | 0.00 [0.00,0.00] |
| **Household density** | | | | |
| Below median | *3988* | 0.00 [0.00,0.00] | 0.00 [0.00,0.00] | 0.00 [0.00,0.00] |
| Above median | *3457* | -0.18 [-0.32,-0.05] | -0.29 [-0.41,-0.16] | -0.30 [-0.43,-0.18] |
| **Household composition** | | | | |
| Alone | *436* | -0.17 [-0.47,0.12] | -0.49 [-0.76,-0.21] | -0.25 [-0.56,0.05] |
| Roomies/friends | *410* | 0.12 [-0.18,0.43] | -0.02 [-0.31,0.26] | 0.14 [-0.17,0.45] |
| Partner | *835* | 0.30 [0.07,0.53] | 0.11 [-0.10,0.33] | 0.23 [0.00,0.46] |
| Parents only | *2884* | 0.00 [0.00,0.00] | 0.00 [0.00,0.00] | 0.00 [0.00,0.00] |
| Parents and siblings/children | *2880* | 0.06 [-0.09,0.21] | 0.11 [-0.03,0.25] | 0.11 [-0.03,0.24] |

CI: Confidence Intervals.
Mean difference and 95 % CI presented.
Adjusted Model 1. Adjusted for age, sex, current education, part-time work, mental well-being at baseline, moving, and geographical region.
Adjusted Model 2. Adjusted model 1, additionally mutually adjusted for other housing conditions

**eTable 2.** Onset of low mental well-being (No. = 6537)

| | | OR (95% CI) | | |
|---|---|---|---|---|
| | *no.* | **Crude Model** | **Adjusted model 1** | **Adjusted model 2** |
| **Direct access to outdoor spaces** | | | | |
| None | *56/246* | 1.97 [1.44,2.70] | 2.17 [1.54,3.05] | 1.68 [1.15,2.47] |
| Common yard | *75/402* | 1.54 [1.18,2.01] | 1.60 [1.20,2.15] | 1.35 [0.98,1.86] |
| Balcony | *64/390* | 1.32 [0.99,1.75] | 1.39 [1.03,1.89] | 1.19 [0.86,1.65] |
| Garden | *499/3842* | 1.00 [1.00,1.00] | 1.00 [1.00,1.00] | 1.00 [1.00,1.00] |
| Multiple outdoor spaces | *218/1657* | 1.01 [0.86,1.20] | 1.07 [0.89,1.27] | 1.00 [0.83,1.21] |
| **Urban** | | | | |
| Urban | *348/2392* | 1.15 [0.98,1.35] | 1.27 [1.05,1.54] | 1.16 [0.95,1.41] |
| Semi-urban | *210/1394* | 1.20 [1.00,1.44] | 1.20 [0.97,1.47] | 1.19 [0.97,1.46] |
| Rural | *354/2751* | 1.00 [1.00,1.00] | 1.00 [1.00,1.00] | 1.00 [1.00,1.00] |
| **Household density** | | | | |
| Below median | *445/3528* | 1.00 [1.00,1.00] | 1.00 [1.00,1.00] | 1.00 [1.00,1.00] |
| Above median | *467/3009* | 1.27 [1.11,1.46] | 1.25 [1.08,1.45] | 1.23 [1.05,1.43] |
| **Household composition** | | | | |
| Alone | *79/369* | 1.70 [1.29,2.23] | 1.83 [1.35,2.47] | 1.47 [1.05,2.07] |
| Roomies/friends | *64/366* | 1.32 [0.98,1.77] | 1.57 [1.14,2.15] | 1.26 [0.88,1.80] |
| Partner | *106/710* | 1.09 [0.86,1.38] | 1.11 [0.86,1.43] | 0.98 [0.74,1.28] |
| Parents only | *351/2535* | 1.00 [1.00,1.00] | 1.00 [1.00,1.00] | 1.00 [1.00,1.00] |
| Parents and siblings/children | *312/2557* | 0.86 [0.73,1.02] | 0.84 [0.71,1.00] | 0.85 [0.72,1.01] |

OR: Odds Ratio, CI: Confidence Intervals.
OR and 95 % CI presented.
Adjusted Model 1. Adjusted for age, sex, current education, part-time work, mental well-being at baseline, moving, and geographical region.
Adjusted Model 2. Adjusted model 1, additionally mutually adjusted for other housing conditions
Participants with low mental well-being scores at baseline excluded (no. = 912).
Mental well-being scores ≤20 points on the SWEMWBS indicate possible or probable depression or anxiety.

**eTable 3.** Changes in QoL (No. = 7445)

| | | Mean differenc (95 % CI) | | |
|---|---|---|---|---|
| | *no.* | **Crude Model** | **Adjusted model 1** | **Adjusted model 2** |
| **Direct access to outdoor spaces** | | | | |
| None | *286* | 0.19 [-0.06,0.45] | -0.19 [-0.43,0.04] | -0.18 [-0.43,0.07] |
| Common yard | *464* | 0.28 [0.08,0.49] | 0.13 [-0.06,0.32] | 0.11 [-0.10,0.31] |
| Balcony | *459* | 0.07 [-0.14,0.28] | -0.09 [-0.28,0.10] | -0.12 [-0.32,0.08] |
| Garden | *4346* | 0.00 [0.00,0.00] | 0.00 [0.00,0.00] | 0.00 [0.00,0.00] |
| Multiple outdoor spaces | *1889* | 0.21 [0.09,0.32] | 0.11 [0.00,0.21] | 0.10 [-0.01,0.20] |
| **Urban** | | | | |
| Urban | *2698* | 0.03 [-0.08,0.14] | -0.06 [-0.17,0.06] | -0.07 [-0.19,0.04] |
| Semi-urban | *1602* | 0.02 [-0.11,0.15] | -0.02 [-0.14,0.10] | -0.01 [-0.13,0.11] |
| Rural | *3145* | 0.00 [0.00,0.00] | 0.00 [0.00,0.00] | 0.00 [0.00,0.00] |
| **Household density** | | | | |
| Below median | *3988* | 0.00 [0.00,0.00] | 0.00 [0.00,0.00] | 0.00 [0.00,0.00] |
| Above median | *3457* | -0.05 [-0.15,0.04] | -0.18 [-0.26,-0.09] | -0.21 [-0.30,-0.12] |
| **Household composition** | | | | |
| Alone | *436* | 0.04 [-0.17,0.26] | -0.26 [-0.45,-0.06] | -0.22 [-0.43,-0.00] |
| Roomies/friends | *410* | 0.08 [-0.14,0.31] | -0.02 [-0.22,0.19] | 0.06 [-0.16,0.29] |
| Partner | *835* | 0.36 [0.20,0.53] | 0.32 [0.17,0.47] | 0.35 [0.18,0.51] |
| Parents only | *2884* | 0.00 [0.00,0.00] | 0.00 [0.00,0.00] | 0.00 [0.00,0.00] |
| Parents and siblings/children | *2880* | -0.06 [-0.17,0.05] | 0.01 [-0.09,0.11] | 0.01 [-0.09,0.10] |

QoL: Quality of Life, CI: Confidence Intervals.
Mean difference and 95 % CI.
Adjusted Model 1. Adjusted for age, sex, current education, part-time work, QoL at baseline, moving, and geographical region.
Adjusted Model 2. Adjusted model 1, additionally mutually adjusted for other housing conditions

**eTable 4.** Onset of low QoL (No. = 6283)

| | | OR (95 % CI) | | |
|---|---|---|---|---|
| | *no.* | **Crude Model** | **Adjusted model 1** | **Adjusted model 2** |
| **Direct access to outdoor spaces** | | | | |
| None | *78/218* | 1.10 [0.83,1.46] | 1.25 [0.92,1.69] | 1.13 [0.81,1.58] |
| Common yard | *113/385* | 0.82 [0.65,1.03] | 0.88 [0.69,1.12] | 0.84 [0.64,1.09] |
| Balcony | *131/379* | 1.04 [0.83,1.30] | 1.10 [0.87,1.39] | 1.06 [0.83,1.37] |
| Garden | *1250/3716* | 1.00 [1.00,1.00] | 1.00 [1.00,1.00] | 1.00 [1.00,1.00] |
| Multiple outdoor spaces | *437/1585* | 0.75 [0.66,0.85] | 0.81 [0.71,0.92] | 0.80 [0.69,0.92] |
| **Urban** | | | | |
| Urban | *706/2273* | 0.93 [0.82,1.05] | 1.02 [0.88,1.18] | 1.03 [0.88,1.19] |
| Semi-urban | *433/1345* | 0.98 [0.85,1.13] | 0.99 [0.84,1.15] | 0.98 [0.84,1.14] |
| Rural | *870/2665* | 1.00 [1.00,1.00] | 1.00 [1.00,1.00] | 1.00 [1.00,1.00] |
| **Household density** | | | | |
| Below median | *1056/3424* | 1.00 [1.00,1.00] | 1.00 [1.00,1.00] | 1.00 [1.00,1.00] |
| Above median | *953/2859* | 1.12 [1.01,1.25] | 1.14 [1.02,1.28] | 1.16 [1.03,1.30] |
| **Household composition** | | | | |
| Alone | *121/338* | 1.17 [0.93,1.49] | 1.37 [1.06,1.78] | 1.38 [1.04,1.83] |
| Roomies/friends | *106/340* | 0.95 [0.75,1.22] | 1.08 [0.83,1.41] | 1.08 [0.81,1.45] |
| Partner | *195/677* | 0.85 [0.71,1.03] | 0.85 [0.69,1.04] | 0.86 [0.70,1.07] |
| Parents only | *786/2442* | 1.00 [1.00,1.00] | 1.00 [1.00,1.00] | 1.00 [1.00,1.00] |
| Parents and siblings/children | *801/2486* | 1.00 [0.89,1.13] | 1.00 [0.88,1.13] | 1.00 [0.88,1.14] |

QoL: Quality of Life, OR: Odds Ratios, CI: Confidence Intervals.
OR and 95 % CI presented.
Adjusted model 1. Adjusted for age, sex, current education, part-time work, QoL at baseline, moving, and geographical region.
Adjusted model 2. Adjusted model 1, additionally mutually adjusted for other housing conditions
Participants with low quality of life at baseline excluded (no. =1162)

**eTable 5.** Onset of loneliness (No. = 6418)

| | | OR (95 % CI) | | |
|---|---|---|---|---|
| | *no. cases/no.* | **Crude Model** | **Adjusted model 1** | **Adjusted model 2** |
| **Direct access to outdoor spaces** | | | | |
| None | *64/234* | 1.56 [1.16,2.11] | 1.82 [1.34,2.48] | 1.44 [1.01,2.04] |
| Common yard | *74/394* | 0.96 [0.74,1.25] | 1.05 [0.80,1.38] | 0.98 [0.73,1.34] |
| Balcony | *89/378* | 1.28 [1.00,1.64] | 1.41 [1.09,1.83] | 1.37 [1.04,1.82] |
| Garden | *736/3793* | 1.00 [1.00,1.00] | 1.00 [1.00,1.00] | 1.00 [1.00,1.00] |
| Multiple outdoor spaces | *295/1619* | 0.93 [0.80,1.07] | 0.99 [0.85,1.15] | 0.97 [0.83,1.13] |
| **Urban** | | | | |
| Urban | *454/2315* | 1.01 [0.88,1.16] | 1.04 [0.88,1.22] | 1.00 [0.85,1.19] |
| Semi-urban | *276/1383* | 1.04 [0.88,1.22] | 1.04 [0.87,1.24] | 1.02 [0.85,1.21] |
| Rural | *528/2720* | 1.00 [1.00,1.00] | 1.00 [1.00,1.00] | 1.00 [1.00,1.00] |
| **Household density** | | | | |
| Below median | *638/3473* | 1.00 [1.00,1.00] | 1.00 [1.00,1.00] | 1.00 [1.00,1.00] |
| Above median | *620/2945* | 1.18 [1.05,1.34] | 1.21 [1.07,1.37] | 1.30 [1.14,1.48] |
| **Household composition** | | | | |
| Alone | *121/362* | 1.94 [1.53,2.46] | 2.37 [1.84,3.06] | 2.12 [1.59,2.82] |
| Roomies/friends | *51/352* | 0.65 [0.48,0.89] | 0.74 [0.54,1.03] | 0.66 [0.47,0.94] |
| Partner | *111/698* | 0.73 [0.58,0.91] | 0.77 [0.61,0.98] | 0.72 [0.56,0.92] |
| Parents only | *514/2498* | 1.00 [1.00,1.00] | 1.00 [1.00,1.00] | 1.00 [1.00,1.00] |
| Parents and siblings/children | *461/2508* | 0.87 [0.76,1.00] | 0.85 [0.74,0.98] | 0.86 [0.74,0.99] |

OR: Odds Ratios, CI: Confidence Intervals.
OR and 95 % CI presented.
Adjusted Model 1. Adjusted for age, sex, current education, part-time work, loneliness at baseline, moving, and geographical region.
Adjusted Model 2. Adjusted model 1, additionally mutually adjusted for other housing conditions
Participants with loneliness at baseline excluded (no. = 1027)

**eTable 6.** Changes in mental well-being, quality of life and loneliness, stratified by sex (No.= 7445)

| | Female | | Male | |
|---|---|---|---|---|
| **Mental well-being** | no. | Mean difference (95 % CI) | no. | Mean difference (95 % CI) |
| **Household density** | | | | |
| Below median | *2816* | 0.00 [0.00,0.00] | *1171* | 0.00 [0.00,0.00] |
| Above median | *2397* | -0.27 [-0.42,-0.12] | *1060* | -0.36 [-0.60,-0.13] |
| **Household composition** | | | | |
| Alone | *278* | -0.02 [-0.39,0.35] | *158* | -0.63 [-1.15,-0.10] |
| Roomies/friends | *267* | 0.40 [0.02,0.78] | *143* | -0.32 [-0.87,0.23] |
| Partner | *655* | 0.24 [-0.02,0.50] | *180* | 0.29 [-0.18,0.76] |
| Parents only | *1971* | 0.00 [0.00,0.00] | *913* | 0.00 [0.00,0.00] |
| Parents and siblings/children | *2042* | 0.21 [0.05,0.38] | *838* | -0.13 [-0.39,0.12] |
| **QoL** | no. | Mean difference (95 % CI) | *no.* | Mean difference (95 % CI) |
| **Household composition** | | | | |
| Alone | *278* | -0.35 [-0.62,-0.08] | *158* | 0.07 [-0.28,0.42] |
| Roomies/friends | *267* | 0.17 [-0.11,0.44] | *143* | -0.11 [-0.48,0.25] |
| Partner | *655* | 0.38 [0.19,0.57] | *180* | 0.19 [-0.12,0.51] |
| Parents only | *1971* | 0.00 [0.00,0.00] | *913* | 0.00 [0.00,0.00] |
| Parents and siblings/children | *2042* | -0.01 [-0.13,0.11] | *838* | 0.05 [-0.12,0.22] |
| **Loneliness**[a] | no. cases/no. | OR (95 % CI) | no. cases/no. | OR (95 % CI) |
| **Household composition** | | | | |
| Alone | *116/278* | 1.81 [1.28,2.56] | *48/158* | 3.15 [1.85,5.36] |
| Roomies/friends | *61/267* | 0.68 [0.45,1.02] | *19/143* | 0.60 [0.28,1.29] |
| Partner | *137/655* | 0.74 [0.56,0.97] | *12/180* | 0.52 [0.26,1.04] |
| Parents only | *563/1971* | 1.00 [1.00,1.00] | *125/913* | 1.00 [1.00,1.00] |
| Parents and siblings/children | *490/2042* | 0.74 [0.62,0.87] | *150/838* | 1.40 [1.04,1.87] |

CI: Confidence Interval, OR: Odds Ratio, QoL: Quality of Life.
Mean difference and 95 % CI and OR and 95 % CI presented.
Results displayed for interaction terms for which the likelihood ratio test indicated interaction (p<0.2).
Adjusted Model 2: Adjusted for age, sex, current education, part-time work, mental well-being/QoL/loneliness at baseline, moving, and geographical region, and additionally mutually adjusted for housing conditions.
[a]Participants with loneliness at baseline excluded (no. = 1023)

**eTable 7.** Joint effects of housing conditions and quarantine on changes in mental well-being, quality of life and loneliness (No. = 7418)

| | | Mean difference (95 % CI) | |
|---|---|---|---|
| | *no.* | **Mental well-being** | **QoL** |
| **Direct access to outdoor spaces** | | | |
| No access, NQ | *b* | -0.98 [-1.38,-0.58] | N/A |
| No access, Q | *b* | -0.48 [-1.12,0.16] | N/A |
| Common yard, NQ | *b* | -0.00 [-0.32,0.32] | N/A |
| Common yard, Q | *b* | -0.57 [-1.08,-0.06] | N/A |
| Balcony, NQ | *b* | 0.02 [-0.29,0.33] | N/A |
| Balcony, Q | *b* | -0.42 [-0.96,0.11] | N/A |
| Garden, NQ | *b* | 0.00 [0.00,0.00] | N/A |
| Garden, Q | *b* | -0.07 [-0.26,0.12] | N/A |
| Multiple outdoor spaces, NQ | *b* | 0.02 [-0.15,0.19] | N/A |
| Multiple outdoor spaces, Q | *b* | -0.13 [-0.40,0.14] | N/A |
| **Urbanity** | | | |
| Urban, NQ | *2073* | N/A | -0.03 [-0.16,0.10] |
| Urban, Q | *615* | N/A | -0.21 [-0.40,-0.03] |
| Semi-urban, NQ | *1223* | N/A | 0.05 [-0.09,0.19] |
| Semi-urban, Q | *373* | N/A | -0.19 [-0.40,0.02] |
| Rural, NQ (referent) | *2355* | N/A | 0.00 [0.00,0.00] |
| Rural, Q | *779* | N/A | 0.01 [-0.14,0.17] |
| **Household density** | | | |
| Below median, NQ (reference) | *3050* | N/A | N/A |
| Below median, Q | *927* | N/A | N/A |
| Above median, NQ | *2601* | N/A | N/A |
| Above median, Q | *840* | N/A | N/A |
| **Household composition** | | | |
| Alone, NQ | *b* | -0.26 [-0.60,0.09] | N/A |
| Alone, Q | *b* | -0.28 [-0.81,0.25] | N/A |
| Roomies/friends, NQ | *b* | 0.05 [-0.30,0.41] | N/A |
| Roomies/friends, Q | *b* | 0.35 [-0.17,0.88] | N/A |
| Partner, NQ | *b* | 0.31 [0.06,0.57] | N/A |
| Partner, Q | *b* | -0.10 [-0.51,0.30] | N/A |
| Parents only, NQ (referent) | *b* | 0.00 [0.00,0.00] | N/A |
| Parents only, Q | *b* | -0.09 [-0.32,0.15] | N/A |
| Parents and siblings/children, NQ | *b* | 0.13 [-0.03,0.29] | N/A |
| Parents and siblings/children, Q | *b* | -0.03 [-0.25,0.20] | N/A |
| **Total** | 7418[a] | | |

CI: Confidence Interval, OR: Odds Ratio, QoL: Quality of Life, Q: Quarantined, NQ: Not Quarantined, N/A: Not applicable.
Mean difference and 95 % CI and OR and 95 % CI presented.
Results displayed for interaction terms for which the likelihood ratio test indicated interaction (p<0.2), otherwise N/A is displayed
Adjusted for age, sex, current education, part-time work, mental well-being/QoL, moving, and geographical region, and additionally mutually adjusted for housing conditions
[a]27 observations omitted due to lacking data on quarantine status
[b]Cells clouded to secure non-identifiability of participants

**eTable 8.** Changes in QoL, stratified by psychiatric illnesses (No. = 7418)[a]

| | No psychiatric illness | | Psychiatric illness | |
|---|---|---|---|---|
| | no. | Mean difference (95 % CI) | *no.* | Mean difference (95 % CI) |
| **Household composition** | | | | |
| Alone | *341* | -0.06 [-0.30,0.18] | *95* | -0.09 [-0.80,0.62] |
| Roomies/friends | *350* | 0.11 [-0.12,0.35] | *60* | -0.40 [-1.26,0.45] |
| Partner | [b] | 0.38 [0.20,0.55] | [b] | 0.46 [-0.17,1.10] |
| Parents only | *2494* | 0.00 [0.00,0.00] | *377* | 0.00 [0.00,0.00] |
| Parents and siblings/children | [b] | -0.01 [-0.11,0.10] | [b] | 0.08 [-0.35,0.50] |
| **Total** | *6450* | | *968* | |

QoL: Quality of Life, CI: Confidence Interval.
Mean difference and 95 % CI presented.
Results displayed for interaction terms for which the likelihood ratio test indicated interaction (p<0.2).
Adjusted Model 2: Adjusted for age, sex, current education, part-time work, QoL at baseline, moving, and geographical region, and additionally mutually adjusted for housing conditions
[a]27 observations omitted due to missing data on psychiatric illnesses.
[b]Cells clouded to secure non-identifiability of participants

**eTable 9.** Changes in category of mental well-being (No. = 7445)

| | no. cases, (ref.normal to normal) low to low/low to normal/normal to low | RRR (95 % CI) Low to low (6.9 %) | Low to normal (5.3 %) | Normal to low (12.3 %) |
|---|---|---|---|---|
| **Outdoor spaces** | | | | |
| No access | (ref. 190)24/16/56 | 1.46 [0.88,2.40] | 1.11 [0.62,1.99] | 1.71 [1.19,2.45] |
| Common yard | (ref. 327)31/32/74 | 1.22 [0.79,1.89] | 1.36 [0.87,2.13] | 1.44 [1.05,1.96] |
| Balcony | (ref. 326)40/29/64 | 1.53 [1.04,2.26] | 1.25 [0.80,1.94] | 1.23 [0.90,1.68] |
| Garden | (ref. 3343)295/209/499 | 1.00 [1.00,1.00] | 1.00 [1.00,1.00] | 1.00 [1.00,1.00] |
| Multiple facilities/other | (ref. 1439)121/111/218 | 1.03 [0.81,1.29] | 1.27 [0.99,1.63] | 1.03 [0.86,1.24] |
| **Urbanicity** | | | | |
| Urban | (ref. 2044)160/146/348 | 0.73 [0.56,0.94] | 0.95 [0.72,1.26] | 1.14 [0.94,1.38] |
| Semi-urban | (ref. 1184)116/92/210 | 0.98 [0.76,1.27] | 1.13 [0.84,1.50] | 1.22 [1.00,1.49] |
| Rural | (ref. 2397)235/159/354 | 1.00 [1.00,1.00] | 1.00 [1.00,1.00] | 1.00 [1.00,1.00] |
| **Household density** | | | | |
| Above median | (ref. 3083)244/216/445 | 1.00 [1.00,1.00] | 1.00 [1.00,1.00] | 1.00 [1.00,1.00] |
| Below median | (ref. 2542)267/181/467 | 1.37 [1.14,1.66] | 1.01 [0.82,1.26] | 1.26 [1.09,1.46] |
| **Household composition** | | | | |
| Alone | (ref. 290)35/32/79 | 1.48 [0.96,2.29] | 1.44 [0.91,2.28] | 1.58 [1.15,2.19] |
| Friends/roommates | (ref. 302)25/19/64 | 1.02 [0.62,1.68] | 0.85 [0.49,1.46] | 1.23 [0.87,1.73] |
| Partner | (ref. 604)64/61/106 | 1.28 [0.92,1.78] | 1.28 [0.90,1.82] | 1.02 [0.78,1.33] |
| Parents only | (ref. 2184)193/156/351 | 1.00 [1.00,1.00] | 1.00 [1.00,1.00] | 1.00 [1.00,1.00] |
| Parents and children/siblings | (ref. 2245)194/129//312 | 0.97 [0.79,1.20] | 0.81 [0.64,1.04] | 0.84 [0.71,0.99] |
| **Total (no.)** | (ref.5625)511/397/912 | | | |

RRR: Relative Risk Ratios, CI: Confidence Intervals.
RRR and 95 % CI presented.
Adjusted Model 2: Adjusted for age, sex, current education, part-time work, moving, geographical region, and mutual adjustment for housing conditions. Mental well-being at baseline omitted from model due to collinearity.

**eTable 10.** Changes in category of QoL (No. = 7445)

| | no. cases, (ref.normal to normal) low to low/low to normal/normal to low | RRR (95 % CI) Low to low (9.5 %) | Low to normal (6.1 %) | Normal to low (27.1 %) |
|---|---|---|---|---|
| **Outdoor spaces** | | | | |
| No access | (ref. 140 )37/31/78 | 1.41 [0.91,2.18] | 1.87 [1.17,3.00] | 1.16 [0.84,1.60] |
| Common yard | (ref. 272)45/35/113 | 0.95 [0.65,1.38] | 1.11 [0.72,1.71] | 0.86 [0.66,1.12] |
| Balcony | (ref. 248)46/34/131 | 1.09 [0.76,1.58] | 1.21 [0.80,1.85] | 1.11 [0.87,1.42] |
| Garden | (ref. 2466)395/235/1250 | 1.00 [1.00,1.00] | 1.00 [1.00,1.00] | 1.00 [1.00,1.00] |
| Multiple facilities/other | (ref. 1148)189/115/437 | 1.06 [0.87,1.30] | 1.03 [0.81,1.32] | 0.81 [0.71,0.93] |
| **Urbanicity** | | | | |
| Urban | (ref. 1567)253/172/706 | 0.88 [0.71,1.10] | 1.05 [0.80,1.37] | 1.04 [0.90,1.21] |
| Semi-urban | (ref. 912)158/99/433 | 0.98 [0.78,1.23] | 1.10 [0.83,1.44] | 0.99 [0.85,1.15] |
| Rural | (ref. 1795)301/179/870 | 1.00 [1.00,1.00] | 1.00 [1.00,1.00] | 1.00 [1.00,1.00] |
| **Household density** | | | | |
| Above median | (ref. 2368)334/230/1056 | 1.00 [1.00,1.00] | 1.00 [1.00,1.00] | 1.00 [1.00,1.00] |
| Below median | (ref. 1906)378/220/953 | 1.51 [1.28,1.79] | 1.12 [0.91,1.37] | 1.18 [1.06,1.32] |
| **Household composition** | | | | |
| Alone | (ref. 217)64/34/121 | 2.14 [1.49,3.08] | 1.49 [0.95,2.35] | 1.39 [1.06,1.83] |
| Friends/roommates | (ref. 234)40/30/106 | 1.15 [0.76,1.73] | 1.38 [0.87,2.20] | 1.05 [0.79,1.40] |
| Partner | (ref. 482)81/77/195 | 1.05 [0.78,1.42] | 1.54 [1.11,2.14] | 0.84 [0.68,1.04] |
| Parents only | (ref. 1656)281/161/786 | 1.00 [1.00,1.00] | 1.00 [1.00,1.00] | 1.00 [1.00,1.00] |
| Parents and children/siblings | (ref. 1685)246/148/801 | 0.84 [0.70,1.01] | 0.90 [0.72,1.14] | 0.98 [0.87,1.11] |
| **Total (no.)** | (ref. 4274)712/450/2009 | | | |

QoL: Quality of Life, RRR: Relative Risk Ratios, CI: Confidence Intervals.
RRR and 95 % CI presented.
Adjusted model 2: Adjusted for age, sex, current education, part-time work, moving, geographical region, and mutual adjustment for housing conditions. QoL at baseline omitted from model due to collinearity.

**eTable 11.** Changes in category of loneliness (No. = 7445)

| | no. cases, (ref.not lonely to not lonely) lonely to lonely/lonely to not lonely/not lonely to lonely | RRR (95 % CI) Lonely to lonely (16.9 %) | Lonely to not lonely (7.6 %) | Not lonely to lonely (6.1 %) |
|---|---|---|---|---|
| **Outdoor spaces** | | | | |
| No access | (ref. 170)26/26/64 | 1.91 [1.16,3.16] | 1.41 [0.87,2.27] | 1.44 [1.02,2.03] |
| Common yard | (ref. 320)39/32/74 | 1.63 [1.08,2.48] | 0.94 [0.62,1.44] | 0.98 [0.73,1.33] |
| Balcony | (ref. 289)28/53/89 | 1.31 [0.84,2.04] | 1.75 [1.23,2.48] | 1.35 [1.02,1.78] |
| Garden | (ref. 3057)255/298/736 | 1.00 [1.00,1.00] | 1.00 [1.00,1.00] | 1.00 [1.00,1.00] |
| Multiple facilities/other | (ref. 1324)115/155/295 | 1.15 [0.90,1.46] | 1.21 [0.98,1.51] | 0.97 [0.83,1.13] |
| **Urbanicity** | | | | |
| Urban | (ref. 1861)169/214/454 | 1.00 [0.77,1.30] | 0.96 [0.76,1.23] | 1.01 [0.85,1.20] |
| Semi-urban | (ref.1107)99/120/276 | 1.01 [0.77,1.33] | 0.98 [0.76,1.26] | 1.02 [0.85,1.21] |
| Rural | (ref.2192)195/230/528 | 1.00 [1.00,1.00] | 1.00 [1.00,1.00] | 1.00 [1.00,1.00] |
| **Household density** | | | | |
| Above median | (ref. 2835)238/277/638 | 1.00 [1.00,1.00] | 1.00 [1.00,1.00] | 1.00 [1.00,1.00] |
| Below median | (ref. 2325)225/287/620 | 1.19 [0.98,1.46] | 1.23 [1.03,1.48] | 1.29 [1.13,1.46] |
| **Household composition** | | | | |
| Alone | (ref. 241)43/31/121 | 1.88 [1.23,2.89] | 1.23 [0.79,1.92] | 2.10 [1.58,2.79] |
| Friends/roommates | (ref. 301)29/29/51 | 1.01 [0.63,1.61] | 0.83 [0.52,1.31] | 0.66 [0.47,0.94] |
| Partner | (ref. 587)38/99/111 | 0.63 [0.42,0.94] | 1.48 [1.11,1.97] | 0.72 [0.56,0.92] |
| Parents only | (ref. 1984)174/212//514 | 1.00 [1.00,1.00] | 1.00 [1.00,1.00] | 1.00 [1.00,1.00] |
| Parents and children/siblings | (ref. 2047)179/193/461 | 0.99 [0.79,1.23] | 0.89 [0.72,1.09] | 0.86 [0.74,0.99] |
| **Total (no.)** | (ref. 5160)463/564/1258 | | | |

RRR: Relative Risk Ratios, CI: Confidence Intervals.
RRR and 95 % CI presented.
Adjusted model 2: Adjusted for age, sex, current education, part-time work, moving, geographical region, and mutual adjustment for housing conditions. Loneliness at baseline omitted from model due to collinearity.

**eFigure 1.** Flow of participation

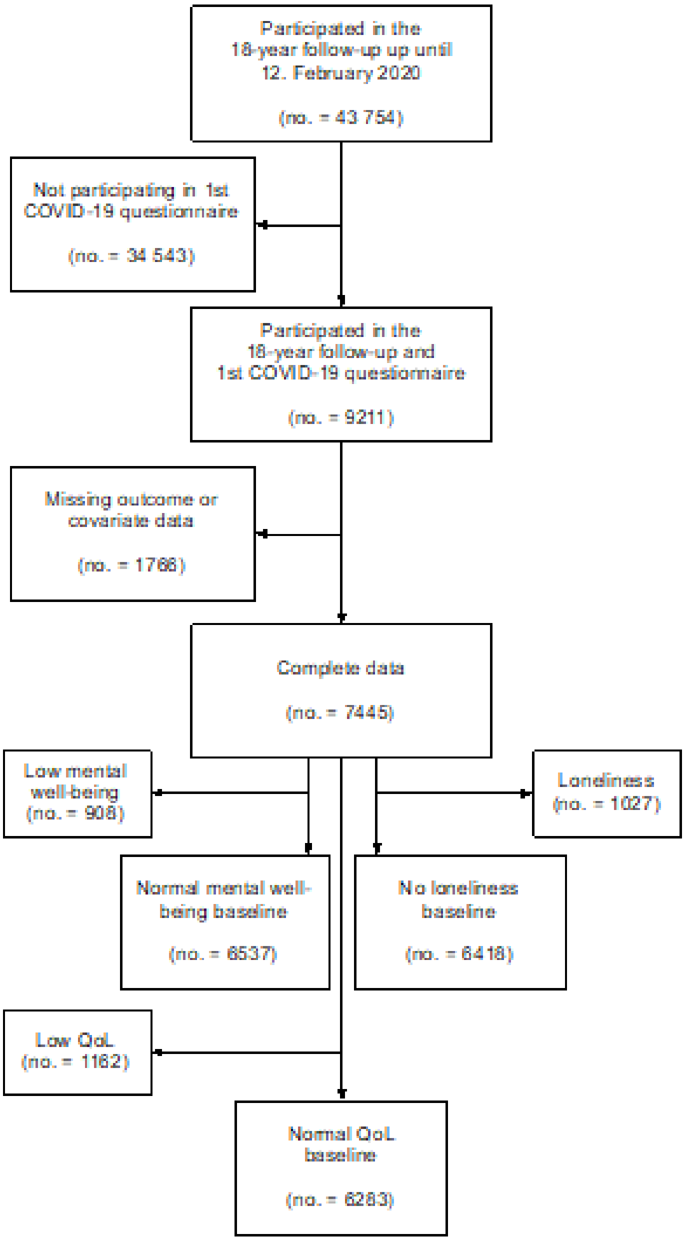
